## Supplemental Figures for "Tracking changes in SARS-CoV-2 transmission with a novel outpatient sentinel surveillance system in Chicago, USA"

**Supplement**

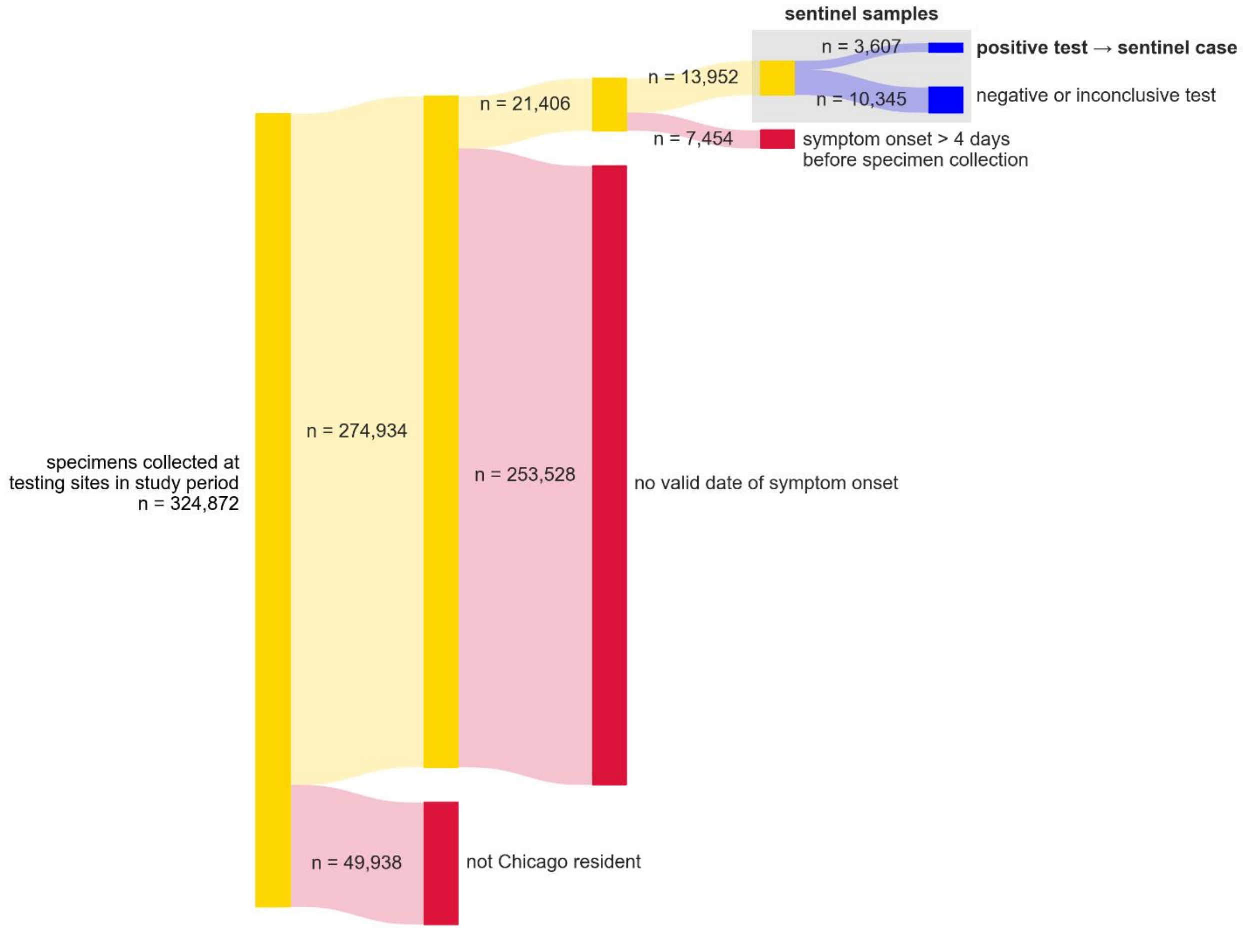


**Figure S1:** Sankey diagram depicting selection of sentinel samples and sentinel cases over the study period (September 27, 2020 to June 13, 2021).

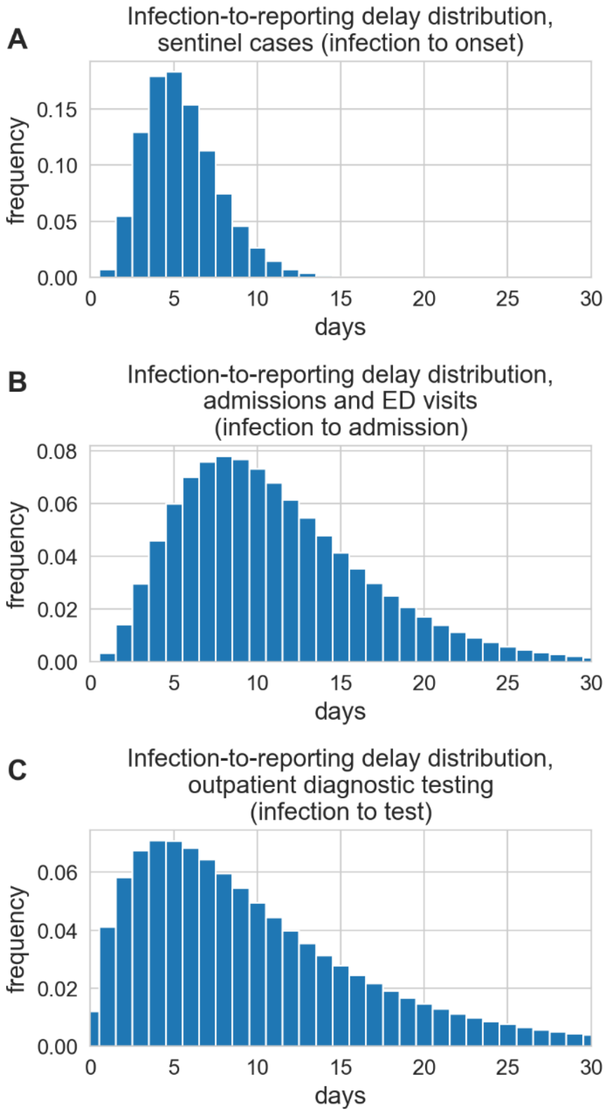


**Figure S2:** Reporting delay distributions used for R(t) estimation. The time from infection to symptom onset **(A)** was approximated with a gamma distribution with shape factor 5.807 and scale factor 0.948 (mean 5.51 days) [21]. The time from infection to hospitalization or emergency department visit **(B)** was approximated with a gamma distribution with shape factor 3.667 and scale factor 3.029 (mean 11.11 days) [18]. The time from infection to test was approximated with epyestim's default reporting delay distribution (mean 10.33 days) [27, 28]. These distributions are based upon research conducted before the global emergence of the Delta variant, which limits their accuracy as the proportion of cases attributable to Delta increased toward the end of the study period, reaching 24% around June 19 2021, after the end of the study.

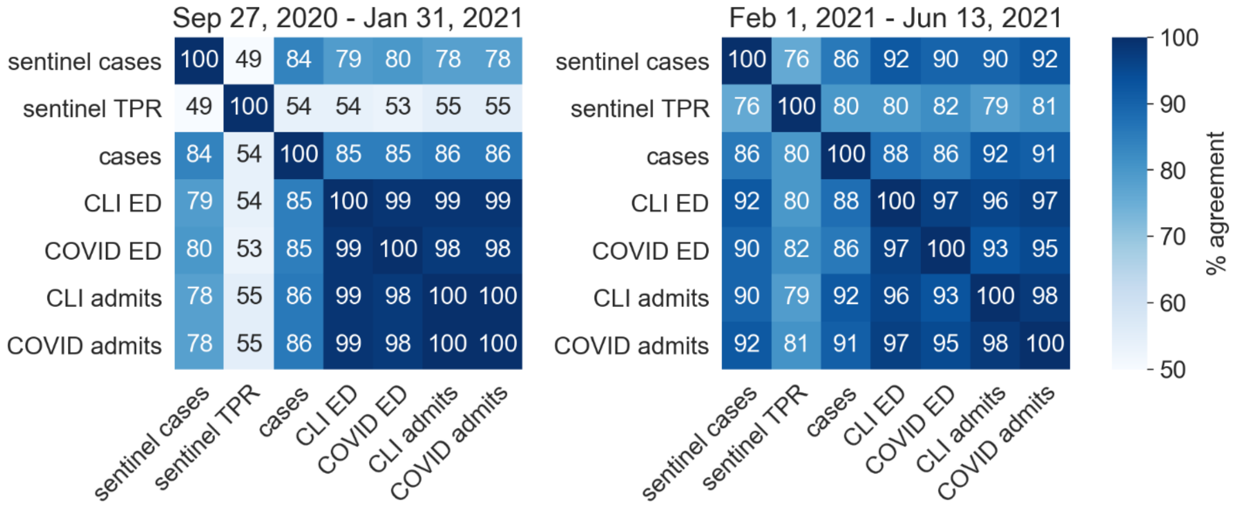


**Figure S3:** Similarity matrices of percent agreement between each pairwise comparison of *R(t)* series for the front half (Sep 27, 2020 – Jan 31, 2021) and back half (Feb 1, 2021 – Jun 13, 2021) of the study window. Percent agreement is the percentage of dates when the median *R(t)* estimates of two series are both ≥ 1.0 or both < 1.0. Compare to **Figure 4B.**

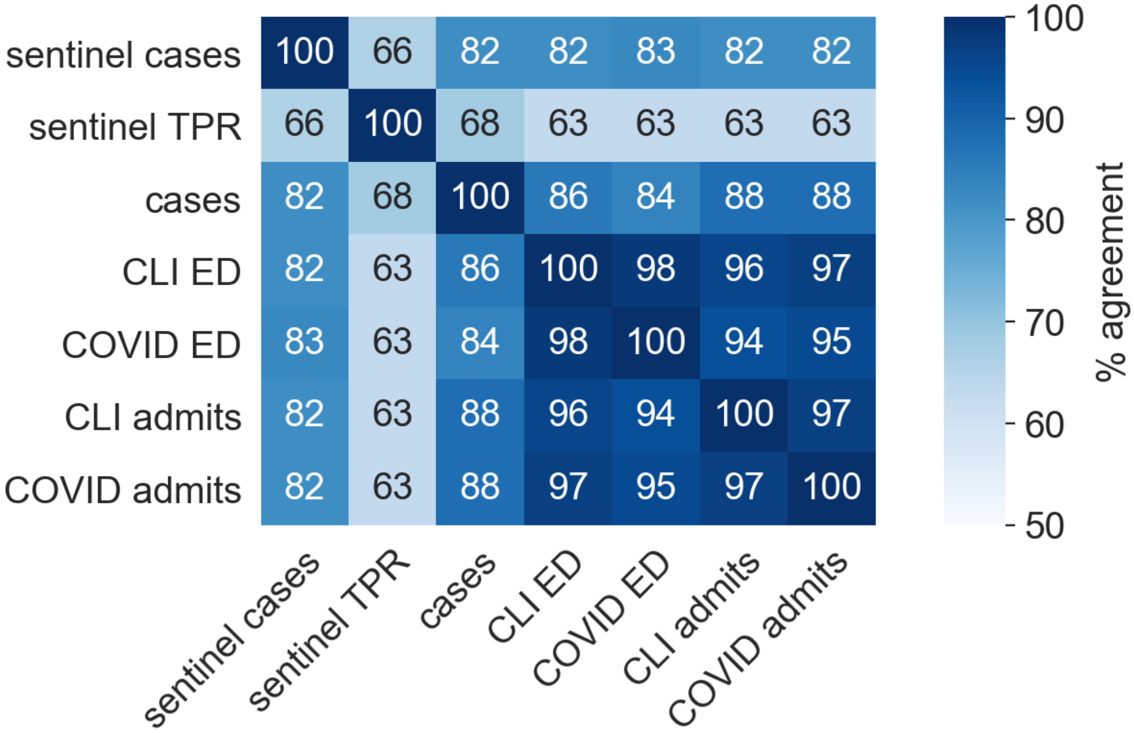


**Figure S4:** Similarity matrix of percent agreement between each pairwise comparison of *R(t)* series, calculated with a seven-day smoothing window (as opposed to a 14-day smoothing window, as presented in **Figure 4B)**. Percent agreement is the percentage of dates when the median *R(t)* estimates of two series are both ≥ 1.0 or both < 1.0.

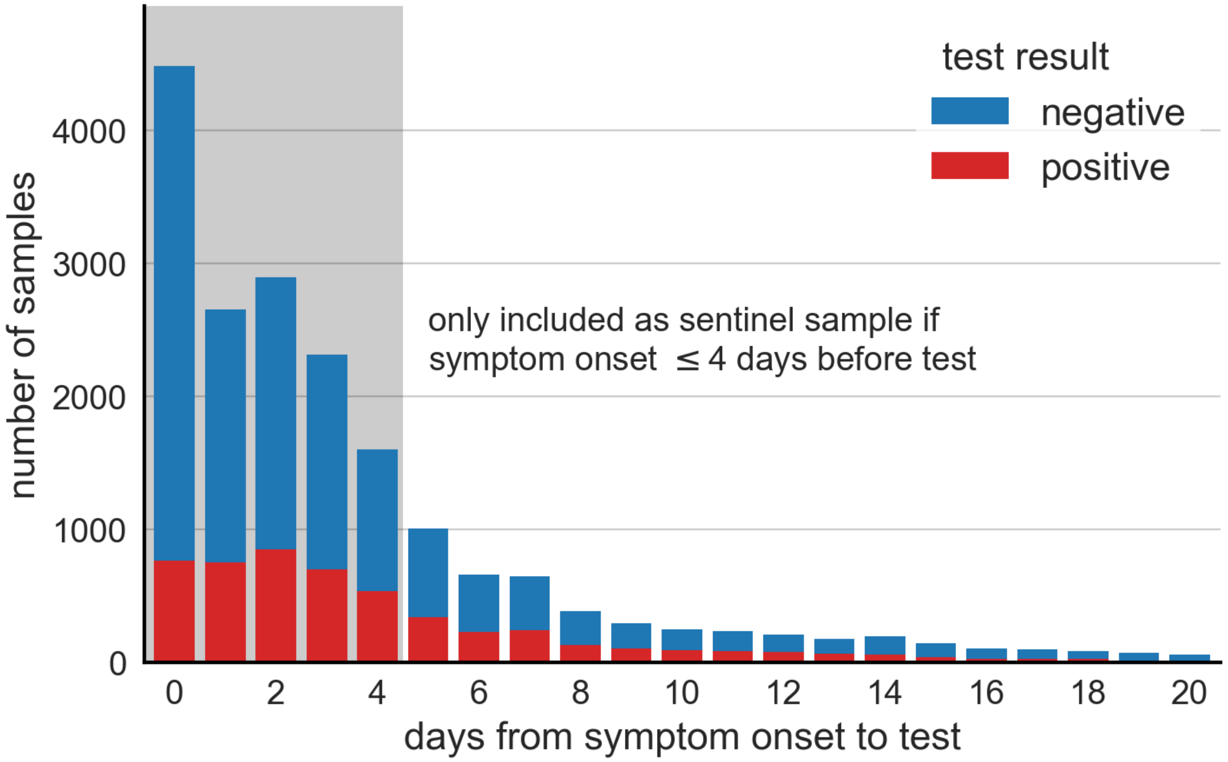


**Figure S5:** Distribution of specimens eligible to be sentinel samples by days elapsed between reported date of symptom onset and date of specimen collection. Time between reported date of symptom onset and specimen collection was greater than 20 days for 2,804 specimens (13.1%).

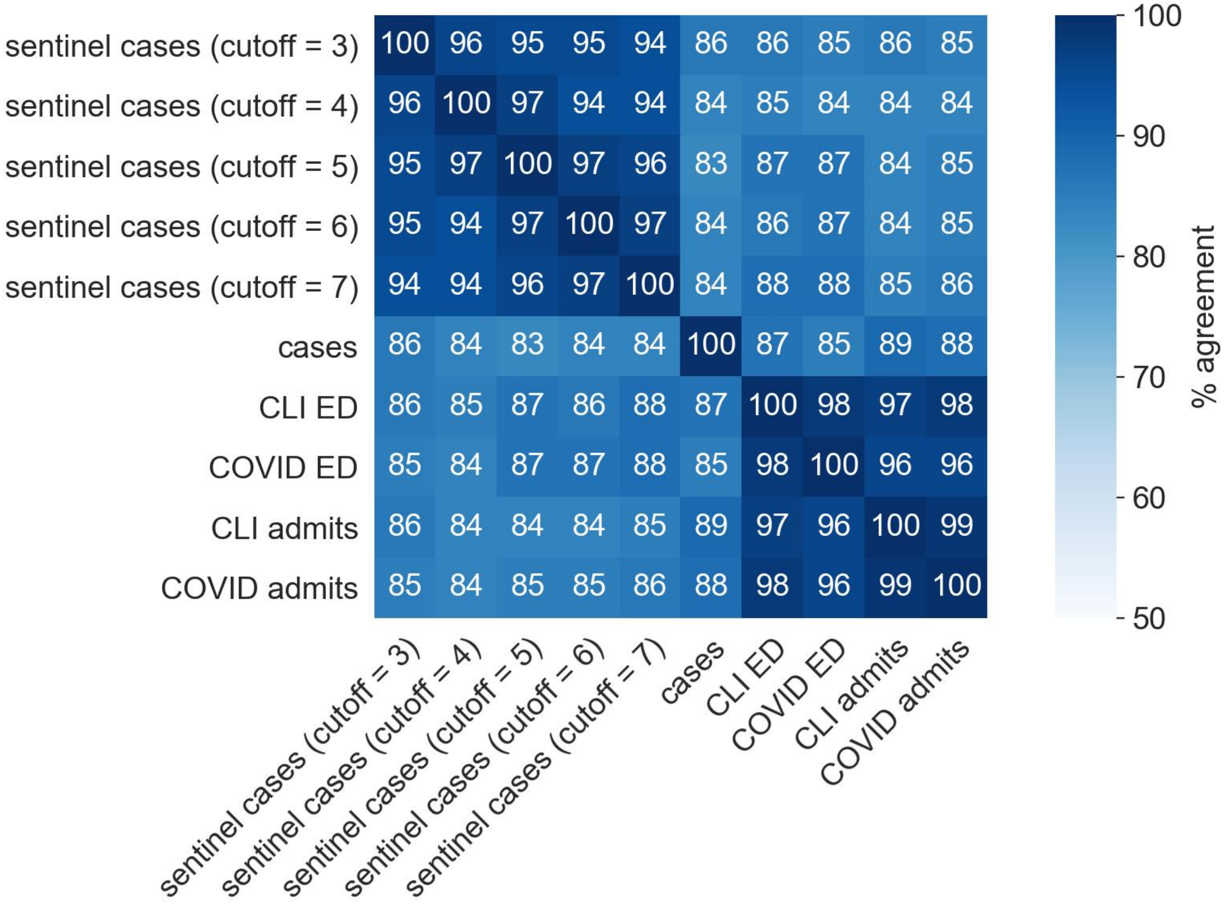


**Figure S6:** Similarity matrix of percent agreement between *R(t)* series. Cutoff indicates the highest allowed value for number of days elapsed between symptom onset and specimen collection in order for a specimen to be considered a sentinel sample. A cutoff value of four days was employed elsewhere in the study. Percent agreement is the percentage of dates when the median *R(t)* estimates of two series are both ≥ 1.0 or both < 1.0.

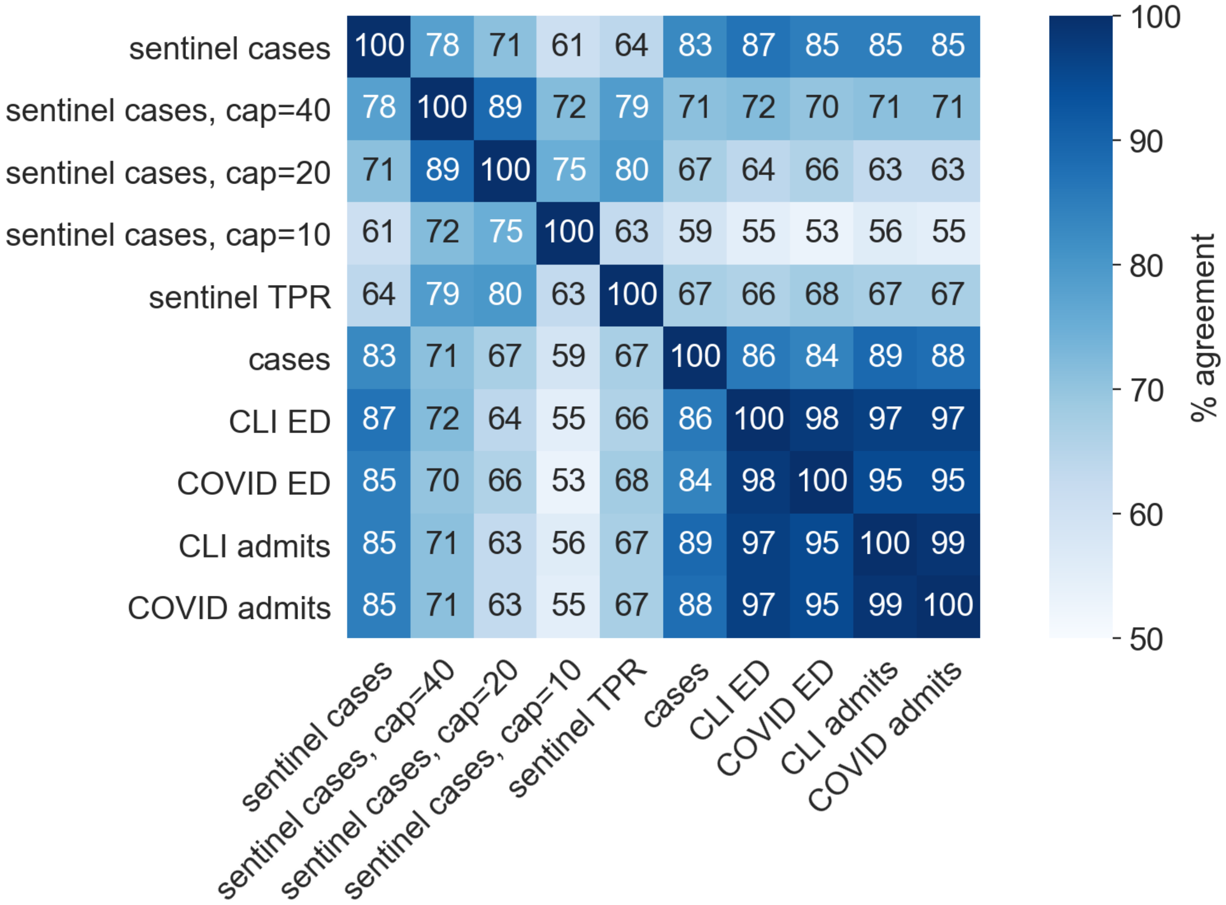


**Figure S7:** Similarity matrix of percent agreement between *R(t)* series. “Cap=X” indicates a subsampling technique wherein only sentinel cases from a random sample of X sentinel samples collected each day were considered. Percent agreement is the percentage of dates when the median *R(t)* estimates of two series are both ≥ 1.0 or both < 1.0.
